# Combined effects of host genetics and diet on human gut microbiota and incident disease in a single population cohort

**DOI:** 10.1101/2020.09.12.20193045

**Authors:** Youwen Qin, Aki S. Havulinna, Yang Liu, Pekka Jousilahti, Scott C. Ritchie, Alex Tokolyi, Jon G. Sanders, Liisa Valsta, Marta Brożyńska, Qiyun Zhu, Anupriya Tripathi, Yoshiki Vazquez-Baeza, Rohit Loomba, Susan Cheng, Mohit Jain, Teemu Niiranen, Leo Lahti, Rob Knight, Veikko Salomaa, Michael Inouye, Guillaume Méric

## Abstract

Co-evolution between humans and the microbial communities colonizing them has resulted in an intimate assembly of thousands of microbial species mutualistically living on and in their body and impacting multiple aspects of host physiology and health. Several studies examining whether human genetic variation can affect gut microbiota suggest a complex combination of environmental and host factors. Here, we leverage a single large-scale population-based cohort of 5,959 genotyped individuals with matched gut microbial shotgun metagenomes, dietary information and health records up to 16 years post-sampling, to characterize human genetic variations associated with microbial abundances, and predict possible causal links with various diseases using Mendelian randomization (MR). Genome-wide association study (GWAS) identified 583 independent SNP-taxon associations at genome-wide significance (*p*<5.0×10^-8^), which included notable strong associations with *LCT* (*p*=5.02×10^-35^), *ABO* (*p*=1.1×10^-12^), and *MED13L* (*p*=1.84×10^-12^). A combination of genetics and dietary habits was shown to strongly shape the abundances of certain key bacterial members of the gut microbiota, and explain their genetic association. Genetic effects from the *LCT* locus on *Bifidobacterium* and three other associated taxa significantly differed according to dairy intake. Variation in mucin-degrading *Faecalicatena lactaris* abundances were associated with *ABO*, highlighting a preferential utilization of secreted A/B/AB-antigens as energy source in the gut, irrespectively of fibre intake. *Enterococcus faecalis* levels showed a robust association with a variant in *MED13L*, with putative links to colorectal cancer. Finally, we identified putative causal relationships between gut microbes and complex diseases using MR, with a predicted effect of *Morganella* on major depressive disorder that was consistent with observational incident disease analysis. Overall, we present striking examples of the intricate relationship between humans and their gut microbial communities, and highlight important health implications.

## Introduction

Humans have co-evolved with the microbial communities that colonize them, resulting in a complex assembly of thousands of microbial species mutualistically living in their gastrointestinal tract. A fine-tuned interplay between microbial and human physiologies can impact multiple aspects of development and health to the point that dysbiosis is often associated with disease^1-3^. As such, increasing evidence points to the influence of human genetic variation on the composition and modulation of their gut microbiota.

Past genetic studies have collectively revealed important host-microbe interactions^4-14^. Previous twin studies detected significant heritability signal from the presence and abundance of only a few microbial taxa, such as some *Firmicutes^15^*, suggesting a strong transientness and variability in gut microbial composition, as well as an important influence from external factors^6,15-18^. Nonetheless, a well-described association between *Bifidobacterium* levels and *LCT-MCM6*, governing the phenotype of lactase persistence throughout adulthood in Europeans, was uncovered in 2015^4^ and subsequently replicated by later studies^6,7,9-12^, suggesting a very strong influence of the evolution of dairy diet in modern humans on their gut bacteria. Additionally, genes involved in immune and metabolic processes^9^ but also disease^19^ were also associated with gut microbial variation. Despite several promising findings, reproducibility across studies varying in sampling and methods is generally poor, and most previously reported associations lose significance after multiple testing corrections^20^. The individual gut microbiota is largely influenced by environmental variables, mostly diet and medication^21-23^, which could explain a larger proportion of microbiome variance than identifiable host genetic factors^9,10^. Biological factors could also influence the cross-study reproducibility of results. GWAS would typically not reproducibly identify genetic associations with taxa harbouring microbial functions potentially shared by multiple unrelated species^24,25^. Indeed, a certain degree of functional redundancy has been observed in human gut microbial communities^25^, which is believed to play a role in the resistance and resilience to perturbations^26-28^. However, both assembly and functioning in human gut microbial communities seem to be driven by the presence of a few particular and identifiable keystone taxa^29^, which exert key ecological and modulatory roles on gut microbial composition independently of their abundance^30,31^. Such taxa are relatively prevalent across individuals and thought to be part of the human “core” microbiota^30,31^, which makes them potentially identifiable through GWAS.

Increasing sample size in studied populations could yield novel and robustly associated results, and alleviate the effect of confounding technical or biological factors. This could be achieved either by performing meta-analyses of GWAS conducted in various populations^12^, or by using larger cohort datasets. In this study, we used a large single homogenous population cohort with matching human genotypes and shotgun faecal metagenomes (N=5959; FINRISK 2002 (FR02)) to identify novel genome-wide associations between human genotypes and gut microbial abundances (**Figure S1**). We further leveraged additional and extensive health registry and dietary individual data to investigate the effects of diet and genotype on particular host-microbial associations, and to predict incident disease linked to gut microbial variation.

## Results

### Genome-wide association analysis of gut microbial taxa

Genome-wide association tests were applied to 2,801 microbial taxa and 7,979,834 human genetic variants from 5,959 individuals enrolled in the FR02 cohort, which includes all taxa discovered to be prevalent in >25% of the cohort (**Methods**). Using a genome-wide significance threshold (*p*<5.0×10^-8^), a total of 478 distinct GTDB taxa, which represented 17% of all tested taxa and included 11 phyla, 19 classes, 24 orders, 63 families, 148 genera and 213 species, were found to be associated with at least one genetic variant (**Figure 1, Table S1**). Conditional analysis found 583 independent SNP-taxon associations at genome-wide significance (**Table S1**). Heritability across the 2,801 taxa ranged between *h^2^*=0.001 to 0.214, with the highest values observed for taxa belonging to the *Firmicutes* and *Firmicutes_A* GTDB phyla, both of which encompassed half (241/476, 50.4%) of all associated taxa with genetic variation (**Figure S2**). There were no differences in SNP heritability between groups of associated or non-associated taxa at genome-wide significance (*p*=0.23).

**Figure 1.**
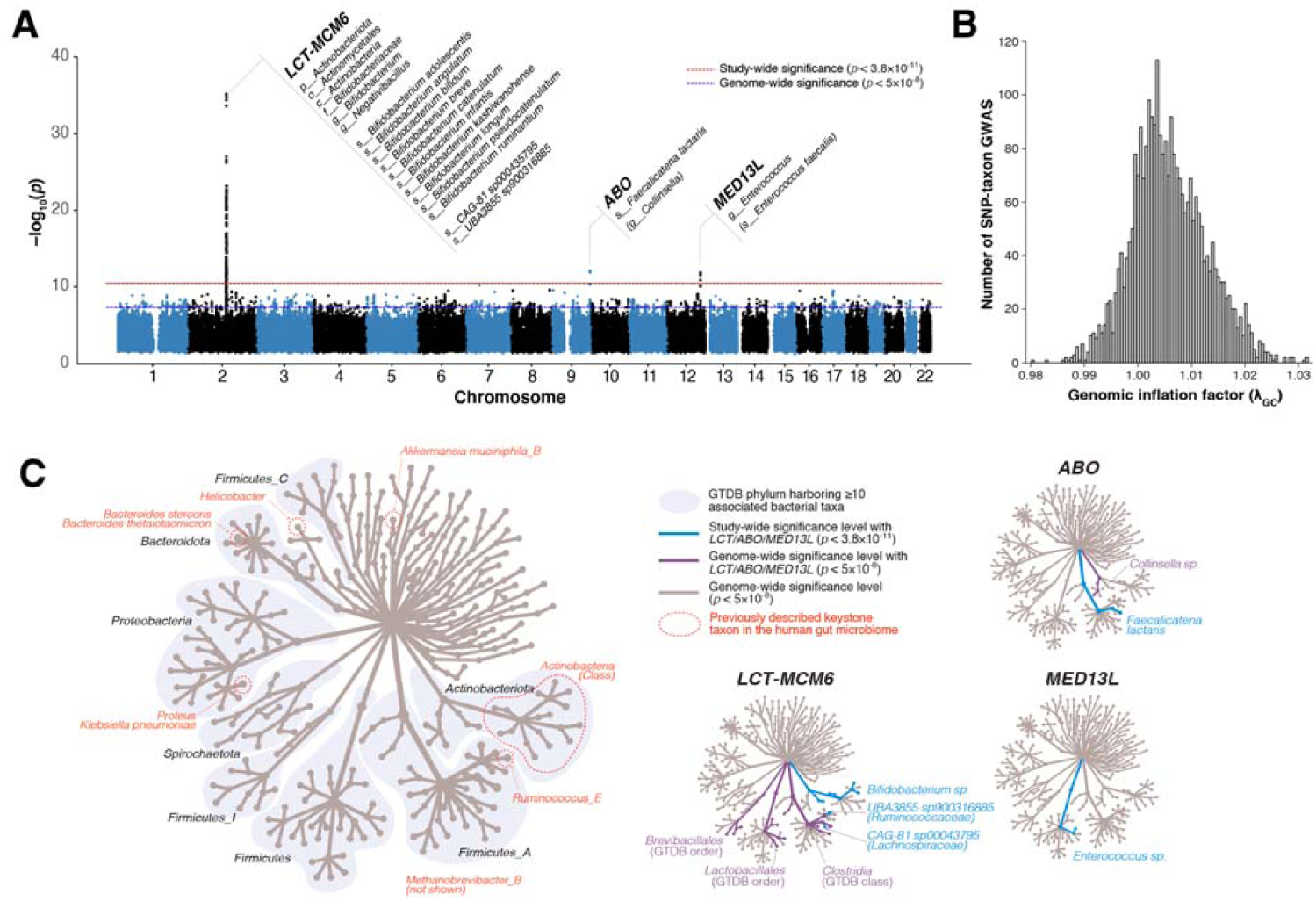
Genome-wide association of human genetic and gut microbial variations. (A) Manhattan plot aggregating the top associations with microbial variation. Each SNP was tested against each of the 2,801 taxa and the Manhattan plot shows the lowest resulting p-value for each SNP. Loci with associations above study-wide significance level (*p*<3.8×10^-11^; red dashed line) are annotated with the human locus name and the corresponding associated microbial taxa. The blue dashed line denotes genome-wide significance level (*p*<5×10^-8^). (B) The distribution of genomic inflation factor (λ_GC_) in 2,801 tested taxa [median(λ_GC_)=1.0051; mean(λ_GC_)=1.0059]. (C) Tree-basec visualization of the taxonomic diversity of genome-wide associated microbial taxa. The central root of the tree represents the Bacteria domain, the first connected node represents phylum, the second connected node class, the third order and the fourth family. Every node represents at least one associated taxa in the GWAS at genome-wide significance level. The three smaller trees on the right highlight all taxonomic groups containing at least one taxon identified as associated with the *LCT-MCM6, ABO*, am: *MED13L* loci (blue edges and nodes denote taxa associated at study-wide significance level and purple edges and nodes denote taxa associated at genome-wide significance level). The main tree is annotated to indicate phyla harbouring >10 distinct genome-wide associated taxa, as well as previously described keystone taxa.

Three loci were strongly associated with microbial variation at study-wide significance, as shown on a Manhattan plot showing the lowest resulting p-value for each SNP tested against each of the 2,801 taxa (**Figure 1, Table 1**). There was no evidence of excess false positive rate in the GWAS (median λ_GC_=1.0051) (**Figure 1B**). After conditional analysis, the strongest association by far (*p*=5.0×10^-35^) involved members of class *Actinobacteria* and rs3940549, a variant in the *LCT-MCM6-ZRANB3* locus region which is in high LD (r^2^=0.87) with the well-described *LCT* variant rs4988235 causing lactase persistence in adults of European ancestry (**Figure S3**). In total, 29 taxa were associated with the *LCT-MCM6* region, including 18 below study-wide significance (**Figure 1, Table S1**). These involved *Bifidobacterium-related Actinobacteriota* and three taxa from the GTDB *Firmicutes_A* phylum which included 2 uncultured species defined from metagenome-assembled reference genomes *(UBA3855 sp900316885* and *CAG-81 sp000435795)* (Table 1). The association of these three *Firmicutes_A* with *LCT* was still genome-wide significant after adjusting for *Bifidobacterium* abundances (**Table S2**). A variant in *ABO* (rs545971), expressing the histo-blood group ABO system transferase, was strongly associated (*p*=1.1×10^-12^) with levels of *Faecalicatena lactaris*. There was evidence for a second independent signal at *ABO* associated with the *Collinsella* genus (chr9:133271182; p=2.5×10^-8^) (**Table S1**, Figure 1). Rs187309577 and rs143507801 in *MED13L*, expressing the Mediator complex subunit 13L, were found to be associated with genus *Enterococcus* (*p*=1.8×10^-12^) and the *Enterococcus faecalis* species (*p*=7.26×10^-11^), respectively (**Table S1**, **Figure 1**).

**Table 1.** Study-wide significant SNP-taxon associations after GWAS. A full table including the associated genotypes and bacterial taxa at genome-wide significance level as well as the full GTDB taxonomic path of all taxa are included in Table S1.

| Locus | Chromosome | Top associated variant (rsID) | Effect allele | Non-effect allele | Associated bacterial taxon (GTDB release 89 nomenclature) | GTDB phylum | If different, best matching taxon synonym(s) from NCBI nomenclature* | beta | SE | P |
| --- | --- | --- | --- | --- | --- | --- | --- | --- | --- | --- |
| LCT-MCM6 | 2 | rs3940549 | G | A | *c_Actinobacteria* | *p_Actinobacteriota* | - | 0.086 | 0.007 | 5.02×10<sup>-35</sup> |
| LCT-MCM6 | 2 | rs3940549 | G | A | *o_Actinomycetales* | *p_Actinobacteriota* | *o_Bifidobacteriales*; *o_Micrococcales* | 0.123 | 0.010 | 3.74×10<sup>-32</sup> |
| LCT-MCM6 | 2 | rs3940549 | G | A | *g_Bifidobacterium* | *p_Actinobacteriota* | - | 0.180 | 0.015 | 1.69×10<sup>-31</sup> |
| LCT-MCM6 | 2 | rs3940549 | G | A | *s_Bifidobacterium_ruminantium* | *p_Actinobacteriota* | - | 0.150 | 0.014 | 2.00×10<sup>-28</sup> |
| LCT-MCM6 | 2 | rs3940549 | G | A | *s_Bifidobacterium_breve* | *p_Actinobacteriota* | - | 0.118 | 0.011 | 8.01×10<sup>-28</sup> |
| LCT-MCM6 | 2 | rs62168795 | C | T | *f_Bifidobacteriaceae* | *p_Actinobacteriota* | - | 0.160 | 0.015 | 8.96×10<sup>-28</sup> |
| LCT-MCM6 | 2 | rs3940549 | G | A | *p_Actinobacteriota* | *p_Actinobacteriota* | *p_Actinobacteria* | 0.051 | 0.005 | 1.20×10<sup>-22</sup> |
| LCT-MCM6 | 2 | rs3940549 | G | A | *s_Bifidobacterium_infantis* | *p_Actinobacteriota* | *s_Bifidobacterium_longum* | 0.112 | 0.011 | 2.37×10<sup>-22</sup> |
| LCT-MCM6 | 2 | rs3940549 | G | A | *s_Bifidobacterium_angulatum* | *p_Actinobacteriota* | - | 0.137 | 0.014 | 4.40×10<sup>-22</sup> |
| LCT-MCM6 | 2 | rs3940549 | G | A | *s_Bifidobacterium_kashiwanohense* | *p_Actinobacteriota* | - | 0.134 | 0.015 | 5.92×10<sup>-20</sup> |
| LCT-MCM6 | 2 | rs182549 | C | T | *s_Bifidobacterium_adolescentis* | *p_Actinobacteriota* | - | 0.207 | 0.023 | 6.11×10<sup>-20</sup> |
| LCT-MCM6 | 2 | rs62168795 | C | T | *s_UBA3855_sp900316885* | *p_Firmicutes_A* | Uncultured *Clostridiales* bacterium | 0.055 | 0.006 | 7.17×10<sup>-19</sup> |
| LCT-MCM6 | 2 | rs3940549 | G | A | *s_Bifidobacterium_pseudocatenulatum* | *p_Actinobacteriota* | - | 0.154 | 0.017 | 8.65×10<sup>-19</sup> |
| LCT-MCM6 | 2 | rs3940549 | G | A | *s_Bifidobacterium_catenulatum* | *p_Actinobacteriota* | - | 0.169 | 0.019 | 1.20×10<sup>-18</sup> |
| LCT-MCM6 | 2 | rs3940549 | G | A | *s_CAG-81_sp000435795* | *p_Firmicutes_A* | Uncultured *g_Clostridium* sp. CAG:58 | -0.151 | 0.017 | 3.72×10<sup>-18</sup> |
| LCT-MCM6 | 2 | rs3940549 | G | A | *s_Bifidobacterium_longum* | *p_Actinobacteriota* | - | 0.131 | 0.015 | 1.41×10<sup>-17</sup> |
| LCT-MCM6 | 2 | rs4988235 | G | A | *s_Bifidobacterium_bifidum* | *p_Actinobacteriota* | - | 0.166 | 0.020 | 1.31×10<sup>-16</sup> |
| ABO | 9 | rs545971 | T | C | *s_Faecalicatena_lactaris* | *p_Firmicutes_A* | *s_Ruminococcus lactaris* (NCBI)<br>*s_Ruminococcus_B lactaris* (GTDB release 86)<br>*s_Mediterraneibacter lactaris* (GTDB release 95) | 0.105 | 0.015 | 1.10×10<sup>-12</sup> |
| MED13L | 12 | rs187309577 | T | C | *g_Enterococcus* | *p_Firmicutes* | - | 0.177 | 0.025 | 1.84×10<sup>-12</sup> |
| LCT-MCM6 | 2 | rs182549 | C | T | *g_Negativibacillus* | *p_Firmicutes_A* | *g_Negativibacillus*; *g_Clostridium* | -0.081 | 0.012 | 5.22×10<sup>-12</sup> |

\* NCBI-GTDB taxonomic synonyms were retrieved using the GTDB Taxon History tool: https://gtdb.ecogenomic.org/taxon_history/

### Human gut microbiome keystone taxa are associated with genetic variation

In total, we identified 31 distinct genetic variants associated (*p*<5.0×10^-8^) with 39 microbial taxa related to identified keystone species as listed by Banerjee *et al*. (20 1 8)^29,32^, which included the *Actinobacteria* class^30^, *Helicobacter pylori*^29^, *Bacteroides stercoris*^33^, *Bacteroides thetaiotaomicron*^34^, *Ruminococcus bromii*^35^, *Klebsiella pneumoniae*^36^, *Proteus mirabilis*^36^, *Akkermansia muciniphila*^31^, and the archaeon *Methanobrevibacter smithii* ^37,38^ (**Figure 1C**, **Table S1**). Only one documented keystone species from Banerjee *et al^29^, Bacteroides fragilis^39^*, was not associated with genetic variation in our study. This observation suggests that keystone species, although defined as exerting selective modulation and not broad effects on microbiome composition variation, generally associates with human genetic variation, suggesting an intimate association with the human gut niche, in line with their reported key ecological roles in microbiome modulation and functioning. Our work highlights novel human genotypes possibly associated with keystone taxa (**Table S1**), which could further improve our understanding of their ecology.

### Combined effect of host genetics and dietary dairy intake on gut levels of Z,C7-associated bacteria

We compared the abundances of 4 bacterial taxa strongly associated with the *LCT* locus *(Bifidobacterium* genus, *Negativibacillus* genus, *UBA3855 sp900316885* and *CAG-81 sp000435795)* in individuals with different rs4988235 genotypes and dairy diets (**Figure 2A**). The abundance of *Bifidobacterium* in individuals producing lactase through adulthood (rs4988235:TT) was unaffected by dairy intake. However, lactose-intolerant individuals (rs4988235:CC) self-reporting a regular dairy diet had a significant increase in *Bifidobacterium* abundance (*p*=1.75×10^-13^; Wilcoxon-rank test). An intermediate genotype (rs4988235:CT) was linked to an intermediate increase (**Figure 2A**). This trend did not seem to be affected by age^40^ (**Figure S4**).

**Figure 2.**
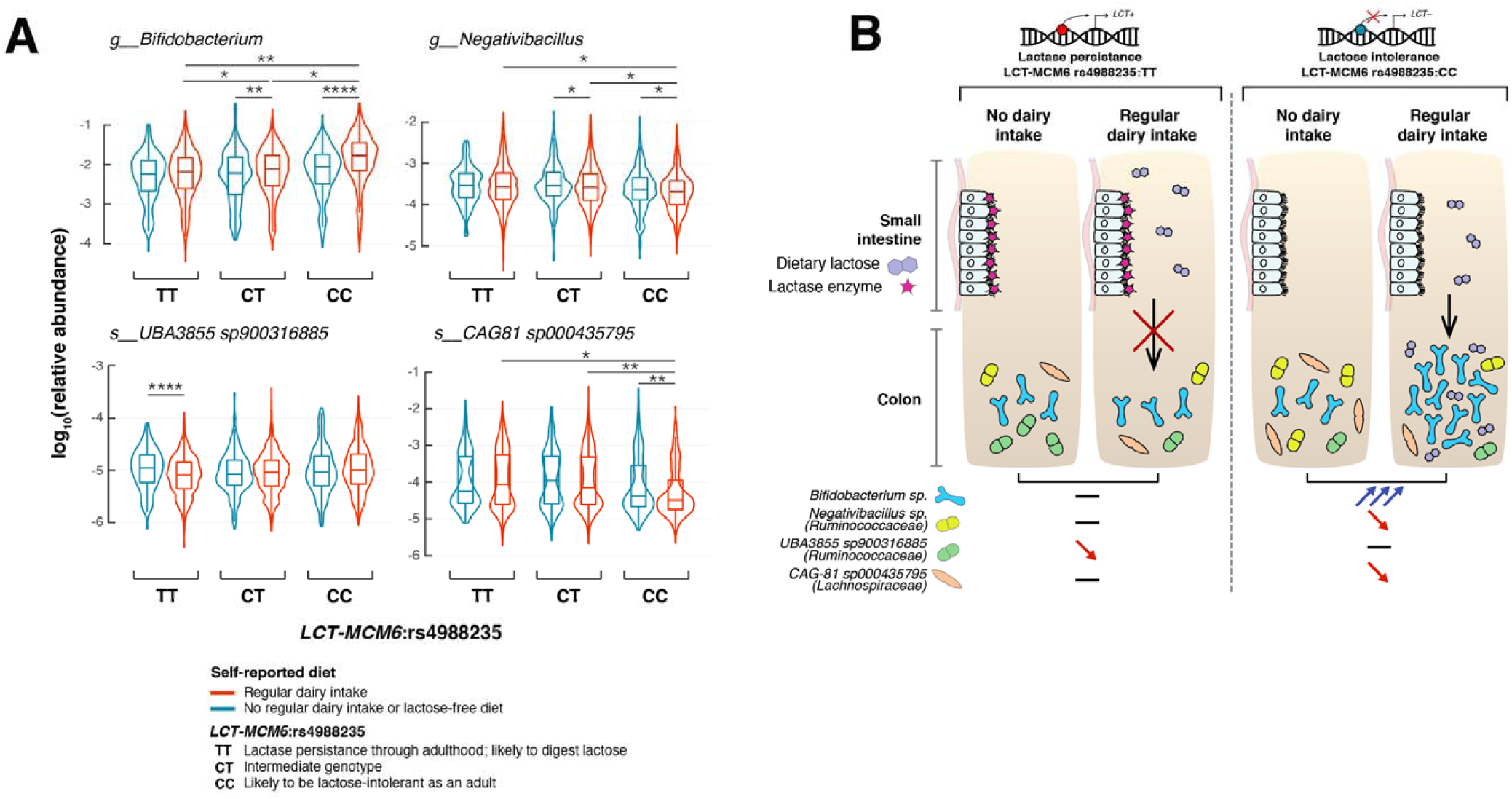
Interaction of human genotype, dairy diet and gut bacterial variation with the *LCT* locus. (A) The 4 panels present variation in microbial abundances for the 4 most significantly associated taxa with the *LCT* locus: *Bifidobacterium, Negativibacillus, UBA3855 sp900316885* and *CAG-81 sp000435795*. Abundances are comparec across stratified groups of individuals from the FR02 cohort according to LCT-MCM6:rs4988235 genotype and self-reported dietary lactose intake (red: regular dairy diet; blue: lactose-free diet). Sample sizes for groups of individuals self-reporting a regular dairy diet: rs4988235:TT (n=1,786), CT (n=2,413), CC (n=736); self-reporting a non-regular dairy diet or lactose-free diet: TT (n=150), CT (n=198), CC (n=245). All statistical comparisons denote the p-values of Wilcoxon rank test on the distributions o! untransformed relative abundances. P-values thresholds are abbreviated as follow: *:*p≤*0.05; **:*p≤*0.01; \*\*\**p≤*50.001; ****:*p≤*0.0001. Only significantly different comparisons are indicated. (B) Host genetics and gut microbes interact in the context of dairy intake and lactose intolerance.

An inverse pattern was observed for the abundance distributions of *Negativibacillus* and uncultured *CAG-81 sp000435795*, for which abundances decreased in lactose intolerant individuals reporting dairy intake, as compared to rs4988235:TT individuals consuming dairy products (*p*=0.049 and p=0.041, respectively) (**Figure 2A**). Levels of *UBA3855 sp900316885* were unaffected by a dairy diet in lactose-intolerant individuals but were surprisingly lower in rs4988235:TT individuals who reported dairy intake (*p*=8.23×10^-5^) (**Figure 2A**). These opposite and contrasting effects of dairy intake on associated bacterial abundances in lactose-intolerant individuals could reflect competition for lactose in the gut. Genus *CAG-81* abundances were the most negatively correlated with those of the other *LCT-*associated taxa (**Figure S5**), which suggests that this competition could be strong and prevalent enough to drive co-association at the *LCT* locus, possibly mediated by lactose intake (**Figure 2B**).

### Functional profiling of CAZymes in 11 *Bifidobacterium* species

Of all 11 *Bifidobacterium* species prevalent enough in our study population to be included in the GWAS, only *B. dentium* was not associated with the LCT locus (*p*=1.70×10^-2^), nor was it co-abundant with any other *Bifidobacterium* species (**Figure S6A**). *B. dentium* has previously been suggested to have different metabolic abilities^41^. A clustering of carbohydrate-active enzymes (CAZyme) profiles from reference genomes of all 11 *Bifidobacterium* species revealed that *B. dentium* clustered apart from the 10 other species, which grouped consistently with their co-abundance patterns (**Figure S6B**). *B. dentium* harboured more genes encoding CAZyme families with preferred fiber/plant-related substrates (GH94, G*h^2^*6, GH53) than other *Bifidobacterium* species, which seemed to harbour more milk oligosaccharide-targeting CAZyme families (GH129, GH112) than *B. dentium* (**Figure S6B**), which could relate to the observed association differences. This suggests that bacterial metabolic abilities can be strong drivers of co-abundance, and of association with human genetic variation.

### Functionally distinct *ABO*-associated bacteria are impacted differently by genotype and dietary fiber intake

A variety of bacteria metabolize blood antigens, with potential applications in synthetic universal donor blood production^42,43^. Gut bacteria are particularly exposed to A- and B-antigens in the gut mucosa of secretor individuals^44^. Our associations of *Faecalicatena lactaris* (*p*=1.10×10^-12^) and *CoUinsella* (*p*=2.59×10^-8^) with *ABO* suggest a possible metabolic link with blood antigens. A comparison of CAZyme profiles across a set of reference genomes revealed 3 CAZymes with blood-related activities in *F. lactaris* (GH110^45^, GH136^46^, CBM32^47^), but none in any of 9 *Collinsella* species (**Figure 3A**). More mucus-targeting and less fiber-degrading enzymes were found in *F. lactaris* than *Collinsella* (**Figure 3A**), suggesting distinct functions in the gut.

**Figure 3.**
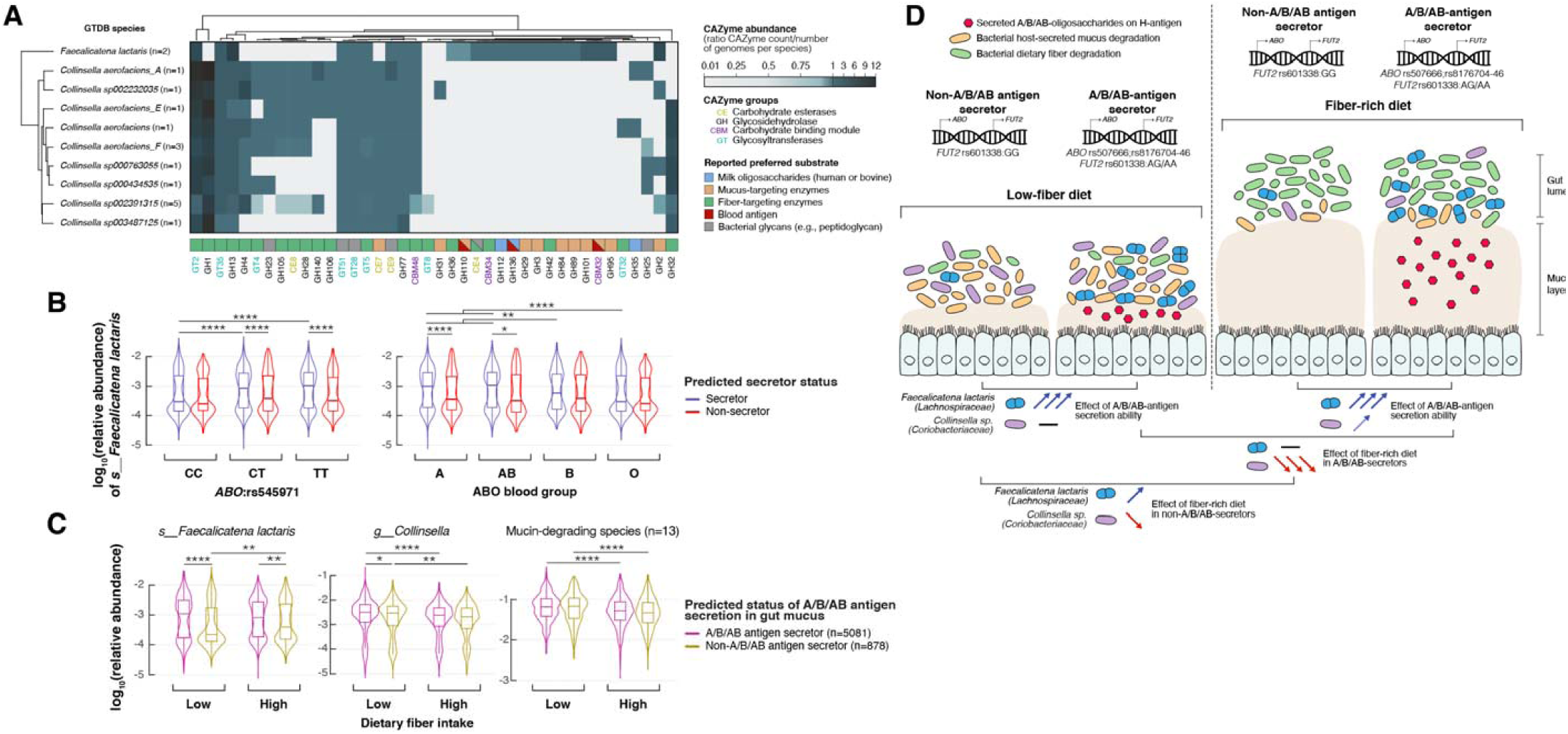
Functional profiling and effect of host genetics and dietary fiber intake on gut abundance variation of two bacterial taxa associated with the *ABO* locus. (A) Carbohydrate-active enzymes (CAZyme) distribution patterns in previously published *F. lactaris* and *Collinsella* reference genomes which were included in the GTDB release 89 index used to classify metagenomes in this study. The heatmap indicates species abundance in corresponding CAZyme families, corresponding to the total count of detected families for each species divided by the number of reference genomes examined for the same species. Values <1 indicate that less than one copy per genome of the corresponding CAZyme family was detected for each, values >1 indicate that more than one copy per genome was detected. Preferred substrate groups are based on literature search and descriptions on CAZypedia.org. (B) ABOassociated *F. lactaris* abundances are compared across stratified groups of individuals from the FR02 cohort according to (left panel): ABO:rs4988235 genotype and predicted secretor status (blue: secretor status conferred by *FUT2* rs601338:AG/AA genotype; red: non-secretor status conferred by *FUT2* rs601338:GG genotype) and (right panel) according to predicted A, AB, B and O blood types, and predicted secretor status. Sample sizes for compared groups of individuals: secretor status with rs545971:C/C (n=1,538), C/T (n=2,493), T/T (n=1,050) and blood group A (n=2,178), AB (n=460), B (n=900), O (n=1,543); non-secretor status with rs545971:C/C (n=266), C/T (n=437), T/T (n=175) and blood group A (n=383), AB (n=80), B (n=148), O (n=267). (C) ABO-associated *F. lactaris* and *Collinsella sp*. abundances, as well as compounded abundances from 13 mucin-degrading species from Tailford *et al*. (2015), are compared across stratified groups of individuals from the FR02 cohort according to the predicted A/B/AB-antigen secretion status and dietary fiber intake. The A/B/AB-antigen secretion status was defined to segregate individuals according to the predicted phenotype of releasing soluble A/B/AB oligosaccharides branched onto a H-antigen into the gut mucosa. A/B/AB-antigen secretors were defined as secretor individuals from blood types A, AB and B. Non- A/B/AB-antigen secretors were defined as non-secretor individuals and O-antigen secretors. Fiber intake was compared in individual groups from the top and bottom quartiles of total fiber score based on dietary questionnaires and approximating the amount of fiber in an individual’s diet. Sample sizes for compared groups of individuals: A/B/AB-antigen secretors (n=1393) following a low-fiber diet (n=723) or a fiber-rich diet (n=670), or non- A/B/AB-antigen secretors (n=952) following a low-fiber diet (n=490) or a fiber-rich diet (n=462). All statistical comparisons denote the p- values of Wilcoxon rank test on the distributions of untransformed relative abundances. P-values thresholds are abbreviated as follow: *:*p≤*0.05; **:*p≤*0.01; ***:*p≤*0.001; ****:*p≤*0.0001. Only significantly different comparisons are indicated. (D) Host genetics and gut microbes interact in the context of fiber intake, secretor status and blood types.

As previously reported^5^, neither ABO blood types, nor secretor status had an impact on alpha and beta diversity (**Figure S7**). However, we observed that the effect of *ABO* genotypes on *F. lactaris* levels, underlying the association, were largely driven by secretor status, with increased abundances in secretor individuals from genotype groups rs545971:CT (*p*=3.6×10^-4^) and rs545971:TT (*p*=9×10^-4^), A (*p*=1.24×10^-5^) and AB blood type groups (*p*=1.24×10^-5^), but not in rs545971:CC genotype (*p*=0.4339), or B and O blood types individuals (**Figure 3B**). Levels in non-secretors did not vary across *ABO* genotypes or blood types (**Figure 3B**). Despite a slight increase in blood type A secretors, *Collinsella* only remained minimally affected by secretor status or blood group (**Figure S8A**). Taken together, this suggests that the secretion of soluble A and B-antigens strongly affects *F. lactaris* in the gut, possibly through reduced opportunity to use them as substrate. Both levels of *F. lactaris* and *Collinsella* were significantly higher when individuals were predicted to secrete A-, B- and AB-antigens in their gut mucosa (*p*<2.2×10^-16^ and p=1.3×10^-8^, respectively) (**Figure S8B**).

A high fiber diet is thought to induce a metabolic transition from mucus-degrading to fiber-degrading activities in the colon, as carbohydrates from fiber are more easily metabolized^48^. The increase in *F. lactaris* abundances in A/B/AB-secretors (defined as secreting A-, B- and AB-antigens) compared to non- A/B/AB-secretors remained strongly significant irrespective of fiber intake (*p*=1.15×10^-9^ in the low-fiber diet group, and p=4.4×10^-3^ in the high-fiber diet group), suggesting that either *F. lactaris* has a strong affinity for secreted A/B/AB-antigens, does not efficiently degrade dietary fiber, or will not easily switch to it as an energy source (**Figure 3C**). *F. lactaris* levels were increased in non-A/B/AB-secretors with a high fiber diet, implying a switch to fiber degradation or interaction with fiber-degrading bacteria (**Figure 3C**). *Collinsella* variation in both A/B/AB-secretors and non-A/B/AB-secretors with high- and low-fiber diets was similar to the compounded abundances of 13 major mucin-degrading species in the human gut^49^, suggesting a similar ecological response in stark contrast with *F. lactaris* (**Figure 3C**, **Figure 3D**).

### *MED13L* association with *Enterococcus faecalis* as a putative link with CRC development

The allele frequency of the *MED13L* rs143507801 variant (A>G), associated with levels of *Enterococcus faecalis* (*p*=7.26×10^-11^), was low (MAF=0.0111), consistent with reported allele frequencies in the gnomAD database^50^. In our study population, 131 individuals carried rs143507801:G allele, 130 being heterozygous (GA) and only one being homozygous (GG). We observed that *E. faecalis* levels were increased in heterozygous rs143507801:GA individuals **(Figure 4)**. *E. faecalis* is a gut commensal, but also an opportunist pathogen believed to play a role in colorectal cancer (CRC) development, possibly through direct damaging of colorectal cells^51-56^. *MED13L* and *MED13* encode for Mediator transcriptional coactivator complex modules associating with RNA polymerase II^57^, and as such specifically interact with cyclin-dependent kinase 8 (CDK8) modules described for their oncogenic activation of transcription during colon tumorigenesis^58^. Consequently, we observed slightly higher levels of *E. faecalis* (*p*=0.014) in 14 individuals enrolled in FR02 who had prevalent CRC at the time of sampling **(Figure 4)**. Groups of individuals segregated by allelic variant and CRC status could not be compared robustly due to small sample size. Taken together, these results suggest a possible link between *E. faecalis* and CRC through the MED13 activation of CDK8 in colorectal tumours, which will need to be investigated further.

**Figure 4.**
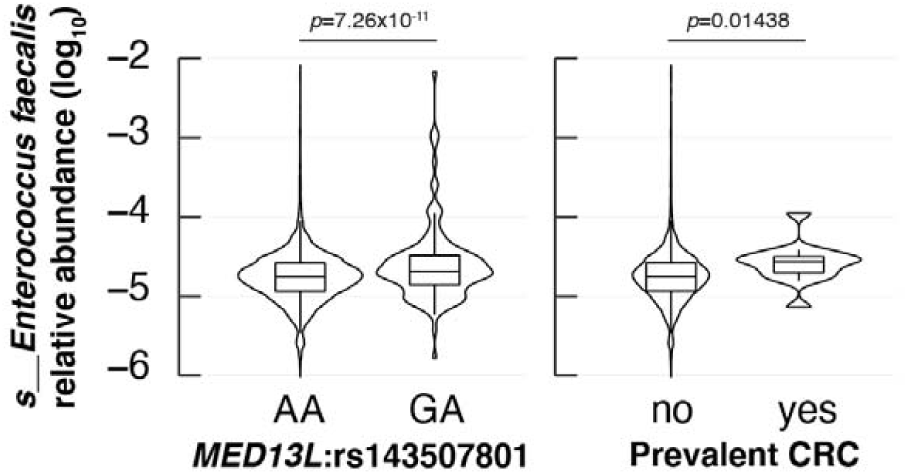
Effect of host genetics and prevalent colorectal cancer on gut levels of *Enterococcus faecalis* associated with *MED13L* variation across participants of the FR02 cohort. Abundances are compared across individuals grouped according to (left panel): MED13L:rs143507801 genotype, (right panel): colorectal cancer (CRC prevalence according to the Finnish Cancer Registry. The comparison between E. *faecalis* variation and MED13L:rs143507801 reflects the GWAS results (Table S1). The comparison of *E. faecalis* abundances in individuals with or without CRC at baseline was performed using a Wilcoxon rank test. Sample sizes for compared groups o] individuals: rsl43507801:A/A (n=5,825), G/A (n=130) (Note: only 1/5959 individual in our cohort was G/G); with CRC (n=14), without CRC at baseline (n=5,941).

### Causal inference predictions between microbes and diseases highlight causal effect of *Morganella* on MDD

Interpreting results of causal inference prediction using bacterial information entails to particular caution, due to the possibility of multiple and unaccounted confounding factors^11^, but can be useful to highlight potential focus for future research. Here, we predicted 96 causal effects in both microbe to disease and disease to microbe directions using bidirectional Mendelian Randomization (MR). Of these, 34 were from microbial levels as exposure to disease as outcome, with a large proportion of causal effects in psychiatric and neurological diseases (**Table S5**). For example, MR suggested an increased abundance of *Faecalicoccus* may have a causal effect on anorexia nervosa (OR=1.8 per SD increase in bacterial abundance; CI95%=1.3-2.5; p=2.0×10^-4^, MR method IVW)(Methods). Other examples included increasing abundances of *Morganella* and *Raoultella* predicted to have causal effects on major depressive disorder (MDD) (**Table S5**). When MR was performed in the reverse direction, using disease risk as an exposure and microbial levels as an outcome, most predicted causal effects involved autoimmune and inflammatory diseases but the strongest predicted causal effect involved type 2 diabetes (T2D) (**Table S6**). Doubling the genetic risk of T2D (possibly accompanied by external factors such as hypoglycaemic medications or metformin intake) was predicted to reduce levels of the uncultured *CAG-345 sp000433315* species *(Firmicutes* phylum) by 0.14 SD (SE=0.04, p=3.0×10^-4^, MR method IVW). A few other examples included some degree of literature validation, such as the higher genetic risk for primary sclerosing cholangitis (PSC) causally impacting levels of the cholesterol-reducing *Eubacterium_R coprostanoligenes^5^*. Furthermore, a higher genetic risk for coeliac disease (CD) was predicted to increase abundances in 4 species previously reported to be more abundant in CD patients than controls^60^ (**Table S6**). Finally, a higher genetic risk for multiple sclerosis (MS) was predicted to cause a reduction in the abundance of *Lactobacillus_B ruminis*, consistent with the report that *Lactobacillus sp*. can reduce symptom severity in an animal model of MS^61^.

The availability in our study dataset of up to 16 years of electronic health record follow-up after the initial sampling of the microbiota allowed for observational validation of predicted effects using MR. Of all causal predictions identified using MR, only the effect of *Morganella* on MDD could be validated by a statistically significant association with incident MDD (HR=1.11, CI_9_5=1.01-1.22, per SD increase of bacterial abundance), after accounting for age, sex and BMI **(Figure 5)**. In our GWAS, *Morganella* variation in the study population associated with a variant (rs 192436108; p=6.16×10^-8^) in the *PDE1A* locus, which has previously been linked to depression^62,63^ and psychiatric disorders^64^. Taken together, these predicted links between *Morganella* and MDD suggest more efforts should be deployed into exploring the possible roles of this bacterium as part of the brain-gut axis metabolic modulation of health.

**Figure 5.**
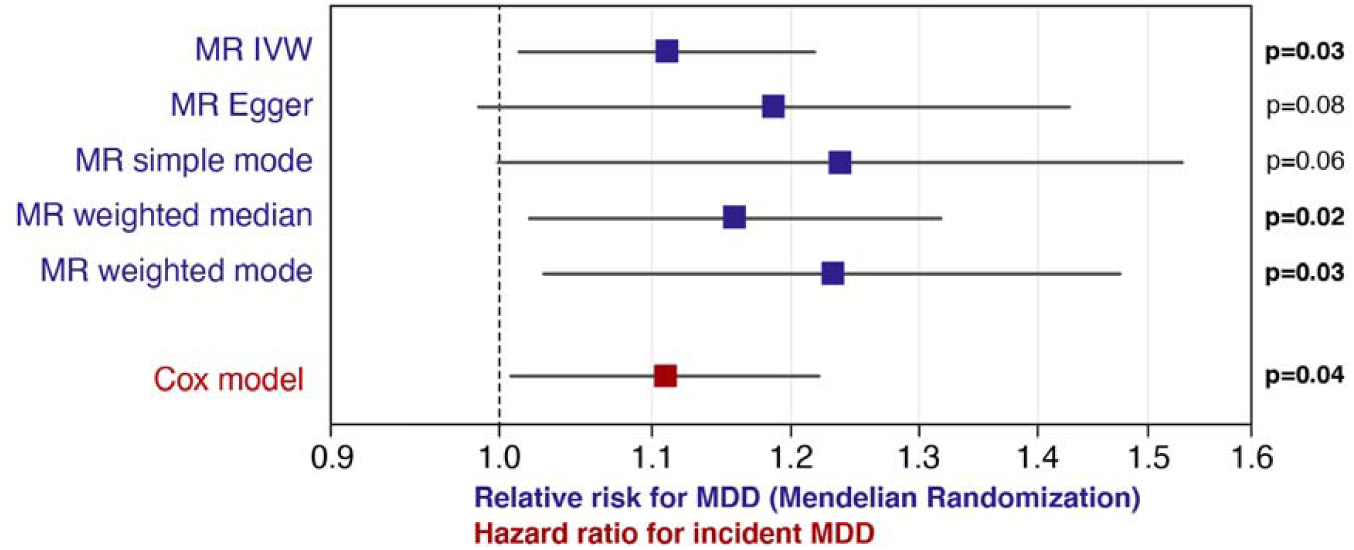
MR-based causal effects and incident depression analysis link *Morganella* with major depressive disorder. The plot shows results for 5 concurring MR methods and hazard ratio for incident MDD in the FR02 cohort up to 16 years after baseline sampling using Cox model.

## Discussion

Here, through GWAS and the subsequent investigation of functional and ecological factors contributing to the most robust human-microbe associations, we present a diverse and global picture of human-microbe interactions in a single cohort of ~6,000 European individuals. We find 3 genetic loci to be strongly associated with gut microbial variation. Two of these loci, *LCT*and *ABO*, are well-known and very segregated in human populations, possibly explaining why our homogenous European cohort identified them as being associated so strongly. A third more mysterious association with the *MED13L* locus highlights possible links with cancer while predictive causal inference highlights several diseases as being causally linked to gut microbes.

### Lactase persistence as a recently evolved strong modulator of gut bacterial abundances

Lactase persistence, or the continued ability to digest lactose into adulthood, is the most strongly selected single-gene trait over the last 10,000 years in multiple human populations^65^, believed to have spread amongst humans with the advent of animal domestication and the culturally transmitted practice of dairying^66^. In our study, as in previous work^4,6,7,11,12^, the association of *LCT* variants with *Actinobacteria*, more specifically *Bifidobacterium*, is by far the most statistically significant, suggesting a profound interaction between *Actinobacteria* and the human gut, in line with their reported keystone activities^30^. We reported a strong increase of *Bifidobacterium* levels in genetically lactose intolerant people reporting a regular consumption of dairy products^9^. This increase was not confounded by age in adults, despite *Bifidobacterium* levels generally decreasing with age in our cohort. While self-reported dietary information is not entirely reliable due to various social reasons^67,68^, our study population was large, and the differences were significant enough to consider this a robust observation. These observations can be explained by the evolutionary adaptation of *Bifidobacterium* species to specifically use human and bovine milk oligosaccharides as an energy source^69^. In adults unable to produce lactase in their small intestines, consumed lactose is likely to become available for colonic bacteria as an energy source to compete for (**Figure 3A**). Hints of a possible competitive relationship between *Bifidobacterium* and *Negativibacillus*, another *LCT-* associated taxon were revealed, which could be mediated by lactose intake and will need to be investigated further in functional studies.

Two interesting questions stem from our findings. First, the genetic determinants of lactose intolerance are known to vary across ethnicity^70^ and cross-population heterogeneity in the *LCT-Bifidobacterium* association was recently reported^12^. As more non-European-centric genetic studies are conducted worldwide^12,71,72^, examining this combined interaction between dairy diet and *Bifidobacterium* in different genetic backgrounds could bring new insights. Secondly, despite recent progresses, lactose intolerance is still largely underdiagnosed, and genetic prediction rates from large population studies exceed lactose intolerance prevalence rates obtained using physical tests^70^. In our work, we lacked information on lactose malabsorption symptoms in lactose intolerant individuals reporting a regular dairy diet. These people could experience discomfort symptoms without knowingly implicating their own lactose intake, but another possibility could be that the ability of *Bifidobacterium* to degrade lactose may alleviate the perceived symptoms of discomfort associated with lactose intolerance, therefore encouraging individuals to unknowingly continue consuming lactose that they would otherwise not be able to digest^73^. This possible probiotic effect would be interesting to investigate in controlled studies.

### Blood antigen secretion can influence levels of specific gut microbial commensals

The *ABO* gene expresses a glycosyltransferase in many cell types, which determines the ABO blood group of an individual by modifying the oligosaccharides on cell surface glycoproteins. A comparison of humans and non-human primates has identified *ABO* (along with the MHC) as harbouring ancient multiallelic polymorphisms that are maintained across species^74,75^. Evolutionary selective pressures at this locus have been proposed to be linked to pathogen infection. Indeed, many infectious diseases such as norovirus infection, bacterial meningitis, malaria, cholera^76^, or even more recently SARS-CoV-2 ^77,78^ are associated with host blood type and secretor status^76^, suggesting that infection could be a driver of a strong balancing selection that has maintained *ABO* polymorphisms. Furthermore, blood type variation has been intriguingly linked to various chronic diseases^76^, such as heart and vascular diseases, gastric cancers, diabetes, asthma or even dementia^76^. Many of these chronic diseases are also associated with dysbiosis of the gut microbiota, which prompts interesting but largely unexplored parallel between gut commensals, blood types and disease^44^. Our study confirms previous findings^5^ that secretor status or blood types do not seem to globally affect gut microbial alpha- or beta-diversity. It also confirms reports from two very recent studies: the first of these studies, a meta-analysis across five German cohorts, using 16S rRNA sequencing to characterize the gut microbiota, linked *Bacteroides* and *Faecalibacterium* to *ABO* and FUT2^79^. The second study, taking a functional approach, intriguingly associated bacterial lactose and galactose degradation genes to *ABO* variation in a cohort of 3,432 Chinese individuals^80^. Taken together, these findings suggest a broad association of *ABO* polymorphisms with microbial variation in various human populations.

An important research effort aiming to enzymatically produce synthetic universal donor blood has driven a push for screening a large diversity of CAZymes, including bacteria, revealing substrate affinities for blood antigens across various microbes^42,43^. Here we highlight *F. lactaris* (formerly *Ruminococcus lactaris)*, as a mucin-degrading commensal likely able to digest blood antigens through its predicted harbouring of GH110, GH136 and CBM32 CAZyme family genes^45-47^. *F. lactaris* is strongly associated with *ABO* genetic variation in our European cohort, and is differentially abundant in people according to their predicted gut mucosal secretion of A/B/AB-antigens. Interestingly, our findings are not consistent with *F. lactaris* switching to a fiber-degrading activity in individuals reporting a high fiber diet, unlike other mucin-degrading bacteria in our study and in the literature^48^ and *Collinsella*, another *ABO-*associated taxon (**Figure 3B**). Our work suggests that some gut commensals such as *F. lactaris* appear to be very efficient and adapted metaboliser of A/B/AB-antigens in the gut, despite their predicted ability to degrade simpler carbohydrates in fiber. This could be an example of ecological niche differentiation in the gut, with impacts on associated *F. lactaris* microbial communities, of which *Collinsella*, also associated with *ABO*, may belong.

### Unexplored links with disease and the nervous system

Although validation of the association is inconclusive because of the low prevalence of CRC cases and genetic variation in our study population, the association of *MED13L* rs143507801 variant with *Enterococcus faecalis* suggested a putative link with CRC. It has been shown that *MED13* could directly link a cyclin-dependent kinase 8 (CDK8) module to Mediator^81,82^, which is a colorectal cancer oncogene, amplified in colorectal tumours and activating transcription driving colon tumorigenesis leading to CRC^58^. This could explain a long suspected link between *Enterococcus faecalis* and development of CRC after having been found in higher concentrations in CRC patients than healthy individuals^51-55^. The suspected mode of action of *E. faecalis* on CRC development is currently unclear, but could be linked to extracellular free radical production directly leading to DNA break, point mutation and chromosomal instability in colorectal cells^56^. Although we saw a trend of *E. faecalis* being increased in abundance in prevalent CRC patients, and in *MED13L* variation, more focused work and a larger sample size will be required to precisely pinpoint a link between this bacterium and CRC through the Mediator complex, if any.

Causal inference analysis highlighted a very promising example of interplay between a gut microbe and a complex disease. Among other suggested links with psychiatric diseases, we predicted that increasing abundances of *Morganella* and *Raoultella* could have causal effects on MDD. Members of the *Enterobacteriaceae* family, such as these two genera, have previously been found in higher levels in MDD patients^83^. Although caution is required when interpreting predictions of causality^84^, several studies elaborated the gut-brain axis hypothesis, and increasing evidence suggests that gut microbes are likely to influence host behavior via a systemic modulation of hormones and metabolites^85-87^. Most importantly, our MR-based observation was consistent with observed hazards using follow-up observational data up to 16 years after initial sampling. This observation supports previous experimental results showing an increase of IgM and IgA-related immune response against *Morganella* secreted lipopolysaccharide in major depression^88^. This finding potentially highlights the intimate influence of the gut-brain axis on humans.

Our MR analysis suggested that known genetic risks of autoimmune and inflammatory diseases could also influence gut microbes. One explanation could be that disease susceptibility would affect host immunity and gut barrier integrity, which may favor an increase in some key microbes. However, several studies have shown that manipulating gut microbial composition could be a potential therapy for autoimmune and inflammatory diseases^89^, which would suggest that composition variation in specific gut microbe maybe a requirement for the penetration of a disease phenotype^90^. Further mechanistic studies are needed to untangle host-microbe interactions in disease, and further interpret these predictions.

### The case for larger datasets and including uncultured novel species in metagenomic studies

Our study highlights the benefits of increasing sample size to increase the statistical power for discovery. Although the *LCT* locus has been reported multiple times to be associated with bacterial taxa, our work is the first to report study-wide significant associations in a single cohort, at the strongest significance ever reported. The association with *Bifidobacterium* in our study was even stronger than the recent findings that used integrative data from 18,473 individuals in 28 different cohorts^12^, emphasizing the importance of standardized methodology and homogeneity in participant ethnicity (especially when studying highly geographically distributed traits such as lactose intolerance traits^91^). *ABO* allelic variation is also notoriously affected by geography^92^, which could explain why some meta-analyses in non-homogenous populations could miss it or not. Importantly, metagenomic sequencing with standardized, robust taxonomic definitions^93,94^ can provide species-level characterization of microbial profiles in the gut of individuals, which is challenging when using 16S rRNA-based studies. An example from our work is the observation that *Bifidobacterium dentium* was prevalent but not associated with the *LCT* locus like all other *Bifidobacterium* species in the population. Observed difference in carbohydrate-active enzymes that are commonly found in other *Bifidobacterium* species may explain this difference^41^. Furthermore, GTDB taxonomic standardization results in greater taxon granularity, i.e. smaller, more discrete clades of similar phylogenetic depth than commonly known lineages or species^93,94^. In theory, this would increase overall accuracy^95^, as a weak association with a poorly-defined lineage may be caused by a strong association with a well-defined subset of that lineage, defined as a coherent group using GTDB^94^. Finally, a myriad of microbial taxa that are to date solely defined and represented by uncultured metagenome-assembled genomes (MAGs) in the GTDB database were found to be independently associated with various loci. Along with recent reports that the more gut microbiome diversity is explored, the more novel, unknown species are discovered^96,97^, this suggests that many discoveries are yet to be made in the field of human microbiome studies.

## Material and methods

### Study population

The FINRISK study population has been extensively described elsewhere^98^. FINRISK population surveys have been performed every 5 years since 1972 to monitor trends in cardiovascular disease risk factors in the Finnish population^98,99^. The FINRISK 2002 (FR02) study population has been extensively described elsewhere^98,100^. Briefly, it was based on a stratified random sample of the Finnish population aged between 25 and 74 years from six geographical areas of Finland^101^. The sampling was stratified by sex, region and 10-year age group so that each stratum had 250 participants. The overall participation rate was 65.5% (n = 8,798). Selected participants filled out a questionnaire, then participated in a clinical examination carried out by specifically trained nurses and gave a blood sample from which various laboratory measurements were performed. They also received a sampling kit and instructions to donate a stool sample at home and mailed it to the Finnish Institute for Health and Welfare in an overnight mail. The follow-up of the cohort took place by record linkage of the study data with the Finnish national electronic health registers (Hospital Discharge Register and Causes of Death Register), which provide in practice 100% coverage of relevant health events in Finnish residents. For present analyses involving follow-up data, we used a follow-up which extended until 31/12/2018.

The study protocol of FR02 was approved by the Coordinating Ethical Committee of the Helsinki and Uusimaa Hospital District (Ref. 558/E3/2001). All participants signed an informed consent. The study was conducted according to the World Medical Association’s Declaration of Helsinki on ethical principles.

### Cohort phenotype metadata and specific dietary information

The phenotype data in this study comprised of demographic characteristics, life habits, disease history, laboratory test results and follow-up electronic health records (EHRs). More specifically, baseline dietary factors were collected. Participants were asked to provide answers to exhaustive diet questionnaires when they were enrolled in the study. Details of the method have been described previously^99^. To broadly assess diet information within the cohort participants, a binary variable was used to indicate whether individuals were self-reporting to follow various possible dietary restrictions. Dietary consumption of specific food product categories was also reported.

### Self-reporting of lactose-free diet and dietary fibre consumption

Allelic distribution at the *LCT-MCM6*:rs4988235 variant responsible for lactase persistence in Europeans was as following in our study population: 1,936 (35%) individuals had the T/T allele conferring a lactase persistence phenotype through adulthood, allowing them to digest lactose, while 981 (18%) individuals had the C/C allele conferring lactose intolerance. Most individuals (n=2,611, 47%) had the intermediate allele C/T making them likely to be able to digest lactose. Most individuals reported a regular dairy intake in their diet (n=5,002, 89%), while 706 (12.5%) individuals reported a regular lactose-free diet.

A total fiber consumption score was calculated from the questionnaires, reflecting the overall consumption of a combination of various fiber-rich foods such as high-fiber bread, vegetables (vegetable foods, fresh and boiled) and berries (fruits, berries and natural juices). The resulting total fiber index values ranged from 9 (low dietary fiber intake) to 48 (high dietary fiber intake), with a median of 33. Comparisons of the effects of low- vs. high-fiber diets were made between the 1^st^ (n=1,213) and 4^th^ (n=1,132) quartiles of the total fiber index.

### Genotyping, imputation and quality control

The genotyping was performed on Illumina genome-wide SNP arrays (the HumanCoreExome BeadChip, the Human610-Quad BeadChip and the HumanOmniExpress) and has been described previously^102^. Stringent criteria were applied to remove samples and variants of low quality. Samples with call rate <95%, sex discrepancies, excess heterozygosity and non-European ancestry were excluded. Variants with call rate <98%, deviation from Hardy-Weinberg Equilibrium (*p*<1×10^-6^), and minor allele count < 3 were filtered. Data was pre-phased by using Eagle2 v2.3^103^. Imputation was performed using IMPUTE2 v2.3.0^104^ with two Finnish-population-specific reference panels: 2,690 high-coverage whole-genome sequencing and 5,092 whole-exome sequencing samples. To evaluate the imputation quality, we compared the sample allele frequencies with reference populations and examined imputation quality (INFO scores) distributions. Imputed SNPs with INFO >0.7 were kept for analysis. Post imputation quality control was carried out by using plink v2.0^105^. Samples with >10% missing rate were removed. Individuals with extreme height or BMI values were further excluded (31 individuals with height<1.47m; 5 with BMI >50 were removed). Both genotyped and imputed SNPs were kept for analysis if they met the following criteria: call rate >90%, no significant deviation from Hardy-Weinberg Equilibrium (*p*>1.0×10^-6^), and minor allele frequency >1%. The post-QC dataset comprised 7,980,477 SNPs.

### Metagenomic sequencing from stool samples

Stool samples were collected by participants and mailed overnight to Finnish Institute for Health and Welfare for storing at −20°C; the samples were sequenced at the University of California San Diego in 2017. The gut microbiome was characterized by shallow shotgun metagenomics sequencing with Illumina HiSeq 4000 Systems. We successfully performed stool shotgun sequencing in n=7,231 individuals. The detailed procedures for DNA extraction, library preparation and sequence processing have been previously described^101^. Adapter and host sequences were removed. To preserve the quality of data while retaining most of the disease cases, samples with a total number of sequenced reads lower than 400,000 were removed.

### Taxonomic profiling, quality filtering and data transformation

Taxonomic profiling of FR02 metagenomes has been described elsewhere^100,106^. Briefly, raw shotgun metagenomic sequencing reads were mapped using the *k*-mer-based metagenomic classification tool Centrifuge^107^ to an index database custom-built to encompass reference genomes that followed the taxonomic nomenclature introduced and updated in the GTDB release 89^93-95^. This implies that unless specified otherwise, all taxonomic names in our study refer to their nomenclature in GTDB, which can be related to the original NCBI nomenclature using the GTDB database server: https://gtdb.ecogenomic.org/taxon_history/.

Gut microbial composition was represented as the relative abundance of taxa. For each metagenome at phylum, class, order, family, genus and species levels, the relative abundance of a taxon was computed as the proportion of reads assigned to the clade rooted at this taxon among total classified reads. The relative abundance of a taxon with no reads assigned in a metagenome was considered as zero in the corresponding profile. For the purpose of this association study and because of reduced accuracy and power when considering rare taxa, we focused on common and relatively abundant microbial taxa, defined as prevalent in >25% studied individuals, and defined with at least 10 mapped reads per individual. For the purpose of association, and as previous studies have reported that only some microbial taxa are inheritable^108^, we also removed taxa with zero SNP-heritability. This filtering resulted in a microbial dataset composed of a total of 2,801 taxa, including 59 phyla, 95 classes, 187 orders, 415 families, 922 genera and 1,123 species.

Taxonomic profiles derived from sequencing data are by nature compositional because of an arbitrary total imposed by the instrument^109^. The compositional data of microbial taxa is not independent and can lead to inappropriate use of linear regression. To overcome this artificial bias, all relative abundance values were transformed by centre-log-ratio (CLR)^110^. CLR transformed data can vary in real space and better fit the normality assumption of linear regression. To minimize the impact of zeros, the reads count profiles were shifted by +1 before the transformation. This process was performed using the R package *compositions*. When visually comparing relative abundances in groups of individuals throughout the manuscript, we used untransformed relative abundances, for better interpretability. Alpha (Shannon index) and beta (Bray-Curtis distance) diversity were calculated at genus level used functions in the R package *vegan*.

### Genome-wide association analysis

The protocol followed in this study was described elsewhere^111^. Briefly, linear mixed model (LMM) implemented in BOLT-LMM^112^ was used to search for genome-wide associations accounting for the individual similarity. Since BOLT-LMM only accepts <1 million SNPs in modelling the genetic relationship matrix, SNPs were pruned at the threshold of r^2^<0.1 (plink2^105^, command *--indep-pairwise 1000 80 0.1)*, resulting in 106,201 independent SNPs. BOLT-LMM automatically performs leave-one-chromosome-out (LOCO) analysis to avoid proximal contamination. Although LMM accounts for the cryptic relatedness in individuals, there are still large population structure cannot be addressed. Thus, the top 10 genetic principal components (calculated by FlashPCA2^113^ based on the pruned SNPs mentioned above) were included as covariates. Age, gender, and genotyping batch were adjusted. As no genetic variant was reported to have large effect size on gut microbiota, statistic estimates were based on infinitesimal model which assumes small non-zero effect for large number of genetic variants. To identify independent associations, GCTA-COJO^114^ was used to conduct approximate conditional and joint analysis using individual genetic data. Window size was set to 10 Mb, assuming SNPs on different chromosomes or more than 10 Mb distance are uncorrelated. The resulting effect size (beta coefficient) indicated the number of standard deviation changes of a taxon’s CLR transformed abundance corresponding to one effective allele increase of SNP.

As microbes interact non-independently with each other in the gut, as part of larger ecological and functional communities, matSpDlite^115,116^ was used to estimate the number of independent tests based on eigenvalue variance, the larger the eigenvalue variance the smaller the number of effective tests. The number of independent tests was 1,328 for 2,801 tested taxa. We used this information to calculate a Bonferroni-adjusted study-wide significant level for significant associations, which was set to 5×10^-8^/1328=3.8×10^-11^. A genome-wide significant threshold was set as 5×10^-8^.

### Prediction of ABO blood groups and secretor status

SNP-based typing of ABO histo-blood group was performed. A combination of four SNPs^117^ was used for the prediction, and a 98% concordance with phenotypically typed ABO histo-blood group has been reported for this method^5^. For blood group allele A, the two different types A1 and A2 were predicted by rs507666 and rs8176704 respectively. Blood group allele B was inferred from rs8176746 and blood group allele O was predicted by rs687289. As the combination of these SNPs are exclusive, no haplotype information was needed. To validate the accuracy of prediction, we compared it with the prediction using a different combination of SNPs^77^. The two predictions were highly consistent, with over 99.9% concordance. In addition, the distribution of ABO groups was consistent with the population distribution found in public database. Secretor status was predicted by the genotype of *FUT2* variant rs601338, where AA or AG genotypes are secretors and GG genotypes are non-secretors. An 100% concordance between the variation in rs601338 and secretor status was reported in a study on Finnish individuals^118^.

### Bidirectional two-sample Mendelian randomization (MR) analysis

Causal relationships between diseases and gut microbiota were investigated at genus and species levels only to maximise interpretability. In total, 213 species and 148 genera associated with at least one variant at genome-wide significant level (*p*<1×10^-8^) were included. GWAS summary results were collected for 46 diseases from MR-Base^119^ (**Table S4**). These included 12 autoimmune or inflammatory diseases, 9 cardiometabolic diseases, 13 psychiatric or neurological diseases, cardiovascular diseases, 4 bone diseases and 8 cancers. For disease with more than one GWAS records, the record with the largest sample size was kept.

Bi-directional causal inference was performed as follows to infer causal effects of microbial abundance variation (exposure) on disease risk (outcome), and of disease (exposure) on microbial abundance levels (outcome). To select the SNP instruments for microbial exposures in our study, we followed recommendations from a previous study showing that associated SNPs below a significance threshold of *p<*1 x10^-5^ had the largest explained variance on microbial features^120^. For each taxon, GCTA-COJO was used to perform a conditional analysis to select independently associated SNPs at *p<*1×10^-5^. SNP instruments for disease exposures were selected at genome-wide significant threshold (*p*<5×10^-8^). Subsequently LD-clumping with a strict threshold (r^2^<0.001 in 1000G EUR within 10 Mb windows) was conducted to select independent instruments with the lowest *p* values for taxa and diseases, respectively.

Effective alleles of all genetic variants were oriented to the risk-increasing alleles of exposures. For each inference, five different MR methods were used to estimate the causal effect: (1) inverse variance weighted (IVW)^121^, (2) weighted median^122^, (3) simple mode^123^, (4) weighted mode^123^ and (5) MR-Egger^124^. IVW is the most sensitive method which requires all instruments are valid. But in reality, it is hard to verify that no any genetic instrument violates any instrumental assumptions. Weighted median only requires at least half of the instruments are valid, making its inference robust to the cases where some instruments violating the assumptions. Simple mode and weighted mode rely on the largest group of similar instruments, reducing the effects of other instruments especially outliers. MR-Egger allows instruments having non-zero pleiotropy and provides way to test and estimate the pleiotropy effect in addition to causal estimate. As these methods are based on different assumptions, the consistency among them indicates a credible estimate^125^, even if discrepancy in these methods does not necessarily suggest the absence of causality. A predicted causal estimate was deemed interesting in our study if: (1) it reached a nominal *p<*0.05 for at least three of the five tested methods (**Table S7**), (2) directionality testing supported the causal direction, and (3) no significant casual effect in the reverse direction. In addition, MR-PRESSO^126^ was used to formally detect and correct for the pleiotropic outliers. Analyses were conducted using the R package *TwoSampleMR*^119^

### Cox proportional hazards regression

Cox proportional hazards regression was conducted to test the association between baseline abundance of gut microbe and incident major depression (16 years follow-up, n=181 incident events). Microbial abundances were CLR-transformed and standardized to zero-mean and unit-variance. The Cox models were stratified by sex and adjusted for age and log-transformed BMI, with time-on-study as the time scale. Participants with prevalent major depression at baseline were excluded. R function *coxph()* in the R package *survival* was used for this analysis.

### Profiling of carbohydrate-active enzymes (CAZymes) in bacterial genomes

The standalone run_dbCAN2 v2.0.11 tool^127^ (https://github.com/linnabrown/run_dbcan) was used to scan for the presence of CAZyme genes from public assembled bacterial genomes taken from the GTDB release 89 reference. We used a CAZyme reference database taken from the CAZy database^128^ (31^st^ July 2019 update). In total, we scanned 327 *Bifidobacterium sp*., 2 *Faecalicatena lactaris* and 15 *Collinsella sp*. reference genomes included in GTDB release 89. Three methods were compared as part of the run_dbCAN2 procedure (HMMER, DIAMOND, and Hotpep). We considered a positive detection result when all three methods agreed on a CAZyme family identification. Identification of preferred reported substrates for the various CAZyme families was done manually from key publications^4, 129^, from literature searches and from the CAZypedia website^130^. Certain CAZyme families have a broad range of substrates, many of which are still unknown, which results in our reported preferred substrates to be as accurate as possible, but non-exhaustive.

### Carbon impact and offsetting

We used GreenAlgorithms v1.0^131^ to estimate that the main computational work in this study had a carbon impact of at least 531.94 kg CO_2_e, corresponding to 560 tree-months. As a commitment to the reduction of carbon emissions associated with computation in research, we consequently funded planting of 30 trees through a local Australian charity, which across their lifetime will sequester a combined estimated 8,040 kg CO_2_e, or 15 times the amount of CO_2_e generated by this study.

## Data Availability

The data for the present study are available with a written application to the THL Biobank as instructed in the website of the Biobank: https://thl.fi/en/web/thl-biobank/for-researchers.

## Acknowledgements

We thank all participants of the FINRISK 2002 survey for their contributions to this work. The FINRISK surveys are mainly funded by budgetary funds from the Finnish Institute for Health and Welfare with additional funding from several domestic foundations. MI was supported by the Munz Chair of Cardiovascular Prediction and Prevention. VS was supported by the Finnish Foundation for Cardiovascular Research. LL was supported by Academy of Finland (decision 295741). ASH was supported by the Academy of Finland, grant no. 321356. RL receives funding support from NIEHS (5P42ES010337), NCATS (5UL1TR001442), NIDDK (U01DK061734, R01DK106419, P30DK120515, R01DK121378, R01DK124318), and DOD PRCRP (W81XWH-18-2-0026). This study was supported by the Victorian Government’s Operational Infrastructure Support (OIS) program, and by core funding from: the UK Medical Research Council (MR/L003120/1), the British Heart Foundation (RG/13/13/30194; RG/18/13/33946) and the National Institute for Health Research [Cambridge Biomedical Research Centre at the Cambridge University Hospitals NHS Foundation Trust] [*]. This work was supported by Health Data Research UK, which is funded by the UK Medical Research Council, Engineering and Physical Sciences Research Council, Economic and Social Research Council, Department of Health and Social Care (England), Chief Scientist Office of the Scottish Government Health and Social Care Directorates, Health and Social Care Research and Development Division (Welsh Government), Public Health Agency (Northern Ireland), British Heart Foundation and Wellcome. *The views expressed are those of the authors and not necessarily those of the NHS, the NIHR or the Department of Health and Social Care.

## Author declaration

The study protocol of FINRISK 2002 was approved by the Coordinating Ethical Committee of the Helsinki and Uusimaa Hospital District (Ref. 558/E3/2001). All participants signed an informed consent. The study was conducted according to the World Medical Association Declaration of Helsinki on ethical principles. All necessary patient/participant consent has been obtained and the appropriate institutional forms have been archived.

